# Ivacaftor, not ivacaftor/lumacaftor, is associated with lower pulmonary inflammation in preschool cystic fibrosis

**DOI:** 10.1101/2022.03.28.22273066

**Authors:** Shivanthan Shanthikumar, Sarath Ranganathan, Melanie R. Neeland

## Abstract

Airway inflammation is a key driver of cystic fibrosis (CF) lung disease. The advent of cystic fibrosis transmembrane conductance regulator (CFTR) modulators has the potential to transform the care of CF, however the direct effect of these therapies on lung inflammation is unknown. Here, we profile airway inflammation in bronchoalveolar lavage (BAL) of preschool aged children with CF on CFTR modulator therapy (ivacaftor or ivacaftor/lumacaftor), untreated children with CF, and age-matched healthy controls. We show that children treated with ivacaftor have lower pulmonary concentrations of inflammatory mediators CCL3, CXCL9, CCL2, IL-8, IL-1β, and IL-6 relative to untreated children with CF. Conversely, there was no significant effect of lumacaftor/ivacaftor treatment on airway inflammation. This is the first work to illustrate a difference in early life airway inflammation with CFTR treatment, highlights the effectiveness of ivacaftor in early life, and suggests that BAL inflammatory profile may represent a biomarker of therapeutic response to treatment.

## MAIN

Airway inflammation in cystic fibrosis (CF) involves an early and non-resolving activation of the innate immune system, characterised by neutrophil infiltration, production of serine proteases such as neutrophil elastase (NE), oxidative stress and high levels of pro-inflammatory cytokines, even in the absence of infection^1^. Currently, there are no effective anti-inflammatory therapies, however significant resources are allocated to identifying such treatments ^1^. Cystic fibrosis transmembrane conductance regulator (CFTR) modulator therapy is transforming the care of CF and is now used in infants as young as 4 months of age. The direct effect of CFTR modulator therapy on lung inflammation is unknown, and it remains unclear whether there remains a role for synergistic anti-inflammatory therapy.

Ivacaftor, the first CFTR modulator, has been shown to lower the levels of NE, IL-8 and IL-1β in the sputum of adults ^2^. A study of sputum of adolescents and adults demonstrated treatment with lumacaftor/ivacaftor was associated with significant reduction in IL-1β but not IL-6, IL-8, TNFα or NE ^3^. One study used peripheral blood monocyte-derived macrophages from adults with CF to show that macrophage phagocytosis was restored to non-CF levels in patients taking ivacaftor alone but not lumacaftor/ivacaftor ^4^. Recently, whole blood transcriptomics revealed that people with CF show widespread overexpression of inflammation-related genes that did not change following treatment with lumacaftor/ivacaftor ^5^. In the only study of lower airway samples from early life, McNally et al ^6^ analysed bronchoalveolar lavage (BAL) obtained from children under 6 years of age, 1 year prior to commencing ivacaftor, immediately prior to treatment, and one year post treatment initiation. Inflammation was compared in children who had samples at similar time points but who were not eligible for ivacaftor treatment. This study showed ivacaftor did not reduce the levels of NE, IL-8 or absolute neutrophil count.

Here, we profile airway inflammation in three clinical groups of children with CF [untreated (n=40), lumacaftor/ivacaftor treated (n=5), and ivacaftor treated (n=9)] and in children without lung disease (children having bronchoscopy for investigation of stridor) (n=8). Untreated children either have a genotype ineligible for modulator therapy, or their parents have elected not to start treatment. All children are enrolled in the AREST-CF cohort, which involves collection of excess BAL, at the time of any clinically indicated procedures for research. Demographics of the study participants are described in Table 1 and Supplementary Table 1. Cytokines were measured in cell-free BAL using the Human Soluble Protein Flex Set Cytometric Bead Array (BD Biosciences) according to the manufacturer’s instructions. Data were acquired on an LSR II X-20 Fortessa and analysed using the BD FCAP Array software. The following 9 inflammatory cytokines and chemokines were assessed: tumour necrosis factor-α (TNFα), chemokine ligand 5 (CCL5), chemokine ligand 3 (CCL3), chemokine (C-X-C motif) ligand 9 (CXCL9), chemokine ligand 2 (CCL2), interleukin-8 (IL-8), interleukin-1β (IL-1β), interferon-α (IFN-α) and interleukin-6 (IL-6). For differential analysis, p-values were determined by Kruskal-Wallis rank sum test and adjusted for multiple hypothesis testing using the Benjamini and Hochberg approach to control the false discovery rate (FDR). FDR-adjusted p<0.05 were considered significant. For cytokines showing an FDR-adjusted significant difference, Dunn’s multiple comparison testing was used to find differences between clinical groups.

**Table 1.**
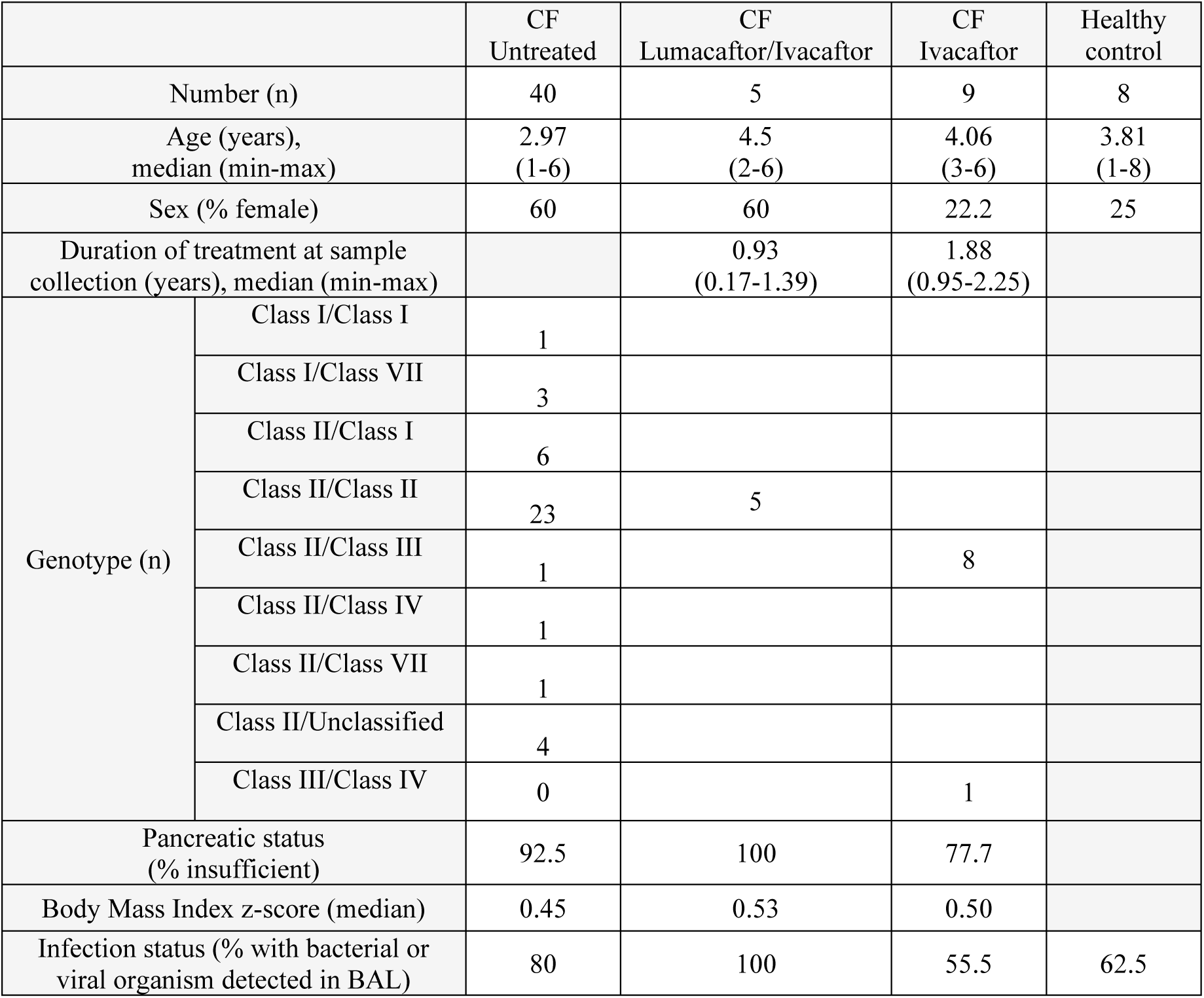
Demographics of study cohort. Genotype was classified based on the system proposed by De Boeck and Amaral ^1^.

|  |  | CF<br>Untreated | CF<br>Lumacaftor/Ivacaftor | CF<br>Ivacaftor | Healthy<br>control |
| --- | --- | --- | --- | --- | --- |
| Number (n) |  | 40 | 5 | 9 | 8 |
| Age (years),<br>median (min-max) |  | 2.97<br>(1-6) | 4.5<br>(2-6) | 4.06<br>(3-6) | 3.81<br>(1-8) |
| Sex (% female) |  | 60 | 60 | 22.2 | 25 |
| Duration of treatment at sample<br>collection (years), median (min-max) |  |  | 0.93<br>(0.17-1.39) | 1.88<br>(0.95-2.25) |  |
| Genotype (n) | Class I/Class I | 1 |  |  |  |
|  | Class I/Class VII | 3 |  |  |  |
|  | Class II/Class I | 6 |  |  |  |
|  | Class II/Class II | 23 | 5 |  |  |
|  | Class II/Class III | 1 |  | 8 |  |
|  | Class II/Class IV | 1 |  |  |  |
|  | Class II/Class VII | 1 |  |  |  |
|  | Class II/Unclassified | 4 |  |  |  |
|  | Class III/Class IV | 0 |  | 1 |  |
| Pancreatic status<br>(% insufficient) |  | 92.5 | 100 | 77.7 |  |
| Body Mass Index z-score (median) |  | 0.45 | 0.53 | 0.50 |  |
| Infection status (% with bacterial or<br>viral organism detected in BAL) |  | 80 | 100 | 55.5 | 62.5 |

Relative to untreated children, ivacaftor-treated children had significantly lower median levels of CCL3 (0.52 vs 4.99 pg/mL, p=0.013), CXCL9 (8.10 vs 34.59 pg/mL, p=0.01), CCL2 (7.61 vs 37.96 pg/mL, p=0.012), IL-8 (21.69 vs 422 pg/mL, p=0.026), IL-1β (1.10 vs 5.93 pg/mL, p=0.010) and IL-6 (0.8 vs 19.89 pg/mL, p=0.001) (Figure 1A). There were no significant differences in BAL cytokines between lumacaftor/ivacaftor treated children and untreated children. Interestingly, median levels of IL-8 and IL-6 were significantly lower in ivacaftor treated children relative to lumacaftor/ivacaftor treated children [IL-8: 21.69 vs 851.48 pg/mL (p=0.03), IL-IL-6: 0.8 vs 47.38 pg/mL (p=0.016)].

**Figure 1.**
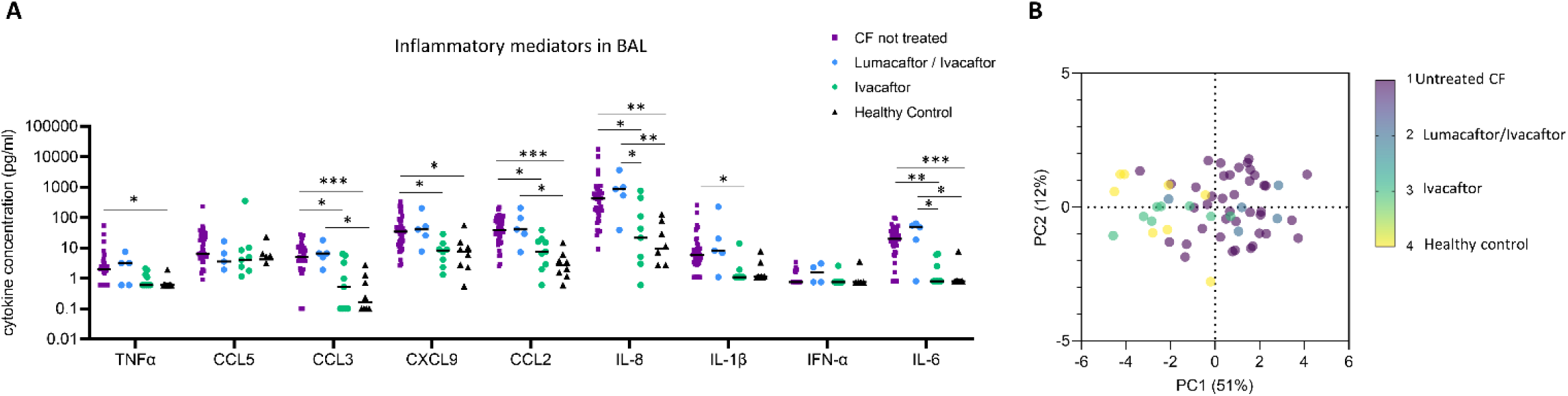
Inflammatory cytokine profile in BAL of children with CF [untreated (purple, n=40), lumacaftor/ivacaftor treated (blue, n=5), and ivacaftor treated (green, n=9)] and healthy controls (black, n=8). **(A)** TNFα, CCL5, CCL3, CXCL9, CCL2, IL-8, IL-1β, IFNα and IL-6 were quantified in BAL using multiplex cytometric bead array. FDR-corrected P values are shown (p<0.05, **p<0.01, ***p<0.001). **(B)** Principal component analysis of all cytokines and samples, untreated CF (purple), lumacaftor/ivacaftor (green), ivacaftor (blue) and healthy controls (yellow). Individual datapoints are shown.

Relative to healthy controls, untreated children with CF showed significantly higher median levels of TNFα (1.97 vs 0.6 pg/mL, p=0.04), CCL3 (4.99 vs 0.16 pg/mL, p=0.0009), CXCL9 (34.59 vs 7.41 pg/mL, p=0.021), CCL2 (37.96 vs 2.72 pg/mL, p=0.0002), IL-8 (422.41 vs 9.38 pg/mL, p=0.001) and IL-6 (19.89 vs 0.8 pg/mL, p=0.001) (Figure 1A). Similarly, lumacaftor/ivacaftor-treated children had significantly higher median levels of CCL3 (6.59 vs 0.16 pg/mL, p=0.036) CCL2 (40.76 vs 2.72 pg/mL, p=0.013), IL-8 (851.48 vs 9.38 pg/mL, p=0.004) and IL-6 (47.38 vs 0.8 pg/mL, p=0.01) relative to healthy controls.

The unsupervised principal component analysis depicted in Figure 1B confirms these findings, with ivacaftor treated children clustering together with the healthy controls, and lumacaftor/ivacaftor treated children clustering together with the untreated CF children.

We demonstrate that children treated with ivacaftor show lower pulmonary concentrations of a range of inflammatory mediators when compared to untreated children, as well as lumacaftor/ivacaftor treated children. In particular, ivacaftor treatment was associated with lower concentrations of two key cytokines: IL-8 and IL-1β which have previously been associated with the development of early life structural lung disease. Treatment with lumacaftor/ivacaftor, however, did not induce reductions of any inflammatory cytokine when compared to untreated children with CF.

Our study also shows lower infection rates in BAL of ivacaftor-treated children relative to lumacaftor/ivacaftor treated children (55.5% vs 100%, p=0.07) (Table 1). Previously ivacaftor was not associated with reduced lower airway infection in preschool children with CF ^6^. However, data from international registries has shown a trend to reduced infections with organisms including *Pseudomonas aeruginosa, Staphylococcus aureus, and Aspergillus species* in people treated with ivacaftor.

There are several limitations of this work which should be highlighted. There are relatively small numbers of patients in the CF treatment groups and participants treated with ivacaftor had been treated for a longer duration than those treated with lumacaftor/ivacaftor (1.88years vs. 0.93 years). In addition, the median age of the untreated CF group was lower than those in the treated group. These limitations reflect the difficulty in obtaining lower airway samples from preschool children. We are only able to obtain samples at the time of clinically indicated procedures which means limits the ability to control duration of therapy and age at sampling. Given highly effective modulator therapy has been shown to improve lung function and sweat chloride after 2 weeks of therapy and this effect is constant to 52 weeks, it is unlikely that longer duration of therapy of lumacaftor/ivacaftor would result in altered inflammatory profile. As the concentration of proinflammatory cytokines increase with age, the younger age of the CF untreated group likely leads to underestimation of the effect of modulator treatment. Despite these limitations, these data still give important insights into the lower airway environment in preschool children with CF and in particular the anti-inflammatory effect of ivacaftor.

While the difference in response to the two treatments is not surprising given prior data showing that ivacaftor is more effective at improving CFTR function and lung disease outcomes, this is the first work to illustrate a difference in airway inflammation in early life. These results suggest that a reduction in early life pulmonary inflammation may be one of the mechanisms by which ivacaftor improves lung disease outcomes. As trials in early life using this drug were predicated on safety and tolerability rather than efficacy, our findings highlight the need for larger, longitudinal studies to determine real-world effectiveness of ivacaftor in optimising lung function (including that determined by more sensitive techniques such as multiple breath washout) and lung structure (preventing the early onset of bronchiectasis). If this was demonstrated with ivacaftor, as well as other novel CFTR modulators such as elexacaftor/tezacaftor/ivacaftor, it would suggest that synergistic anti-inflammatory therapy may not be needed and that BAL inflammatory profile may represent a biomarker of therapeutic response.

## Data Availability

All data produced in the present study are available upon reasonable request to the authors

## Conflict of interest statement

SS, SCR and MRN have no conflicts of interest to declare

## Author Contribution statement

SS and MRN conducted the experiments and co-wrote the first draft of the manuscript. SS and SCR recruited the patients and collected the biospecimens. All authors contributed to and approved the final version of the manuscript.

## Acknowledgements

MRN is supported by a Melbourne Children’s LifeCourse Fellowship provided by the Royal Children’s Hospital Foundation. SS is supported by a Vertex Mentored Research Innovation Award. We thank the children and parents who participated in the AREST-CF study.

## Notes

### Competing Interest Statement

The authors have declared no competing interest.

### Funding Statement

AREST CF is supported by several funding bodies, including the National Health and Medical Research Council and Cystic Fibrosis Australia. SS is supported by a Vertex Mentored Research Innovation Award. MRN is supported by funding from the Chan Zuckerberg Initiative.

### Author Declarations

Ethics approval to conduct this study was obtained from the Royal Childrens Hospital Research Ethics Committee. Written and informed consent was obtained from parents or legal guardians of participants in this study.

